# Comparison of the performance of two targeted metagenomic virus capture probe-based methods using synthetic viral sequences and clinical samples

**DOI:** 10.1101/2023.08.23.23294459

**Authors:** Kees Mourik, Igor Sidorov, Ellen C. Carbo, David van der Meer, Arnoud Boot, Aloysius C. M. Kroes, Eric C.J. Claas, Stefan A. Boers, Jutte J.C. de Vries

## Abstract

Viral enrichment by probe hybridization has been reported to significantly increase the sensitivity of viral metagenomics.

This study compares the analytical performance of two targeted metagenomic virus capture probe-based methods: i) SeqCap EZ HyperCap by Roche (ViroCap) and ii) Twist Comprehensive Viral Research Panel workflow, for diagnostic use. Sensitivity, specificity, limit of detection, and effect of human background DNA were analysed, using synthetic viral sequences, clinical and reference samples with known viral loads.

Sensitivity and specificity were 95% and higher for both methods. Combining thresholds for viral sequence read counts and genome coverage (respectively 500 reads per million and 10% coverage) resulted in optimal prediction of true positive results. Limits of detection were approximately 50-500 copies/ml for both methods. Increasing proportions of spike-in cell free human background sequences did not negatively affect viral detection.

These data show analytical performances in ranges applicable to clinical samples, for both probe hybridization metagenomic approaches. This study supports further steps towards more widespread use of viral metagenomics for pathogen detection, in clinical and surveillance settings using low biomass samples.

## Introduction

Viral metagenomics has been gradually applied for broad-spectrum pathogen detection of infectious diseases^1-5^, surveillance of emerging diseases^3,6-8^, and pathogen discovery^9,10^. Though metagenomic approaches have been practiced for decades in the field of marine environments and the human microbiome, this approach is nowadays changing how physicians diagnose infectious diseases^2^. Whereas amplicon based metagenomics has been successfully adopted for bacterial diagnostics, no amplicon based pan-viral approach is available and therefore viral metagenomics has not yet been widely deployed as diagnostic tool in clinical laboratories^1^. One of the main challenges for application in clinical settings is the low level of viral genomes in the presence of high levels of host material in patient samples. Several methods for depletion of host sequences and enrichment of viral sequences have been studied with varying success rates^11^. For example, depletion of host cells prior to extraction of nucleic acids (NA) has been reported to be not advantageous in clinical samples as also intracellular viral particles or NA will be removed^11,12^. In contrast, viral enrichment by probe hybridization methods has been reported to significantly increase sensitivity in various sample types^1,13-19^, up to the level required for accurate detection of low frequency virus variants^20^.

Previously, the performance of a hybridization capture probe panel targeting vertebrate viruses in cerebrospinal fluids from patients with meningo-encephalitis has been analysed^21^. Viral target sequence read counts increased 100-10.000 fold compared to unenriched metagenomic sequencing, and sensitivity by enrichment was comparable with polymerase chain reaction (PCR)^21^. Moreover, these earlier data showed that this hybridisation panel of approximately two million capture probes designed in 2015 was suited for the detection of novel coronaviruses by reactivity with other vertebrate betacoronavirus probes^10^. During the past years, this specific hybridisation panel distributed by Roche has been adopted in a broad range of different clinical^15,22-29^ and zoonotic settings^22,29,30^. Recently, Twist Bioscience has released a new hybridisation panel containing approximately one million size capture probes targeting human and animal viruses. Reports comparing the performance of viral metagenomic hybridisation panels are lacking.

Here, we compare the analytical performance of two targeted metagenomic virus capture probe-based methods: i) SeqCap EZ HyperCap by Roche (ViroCap) and ii) Twist Comprehensive Viral Research Panel workflow, for clinical diagnostic use. Sensitivity, specificity, and limit of detection were analysed using synthetic viral sequences, clinical and reference samples with known viral loads.

## Methods

### Synthetic sequences and clinical samples

An overview of the validation panels and study design is shown in **Figure 1**.

**Fig. 1.**
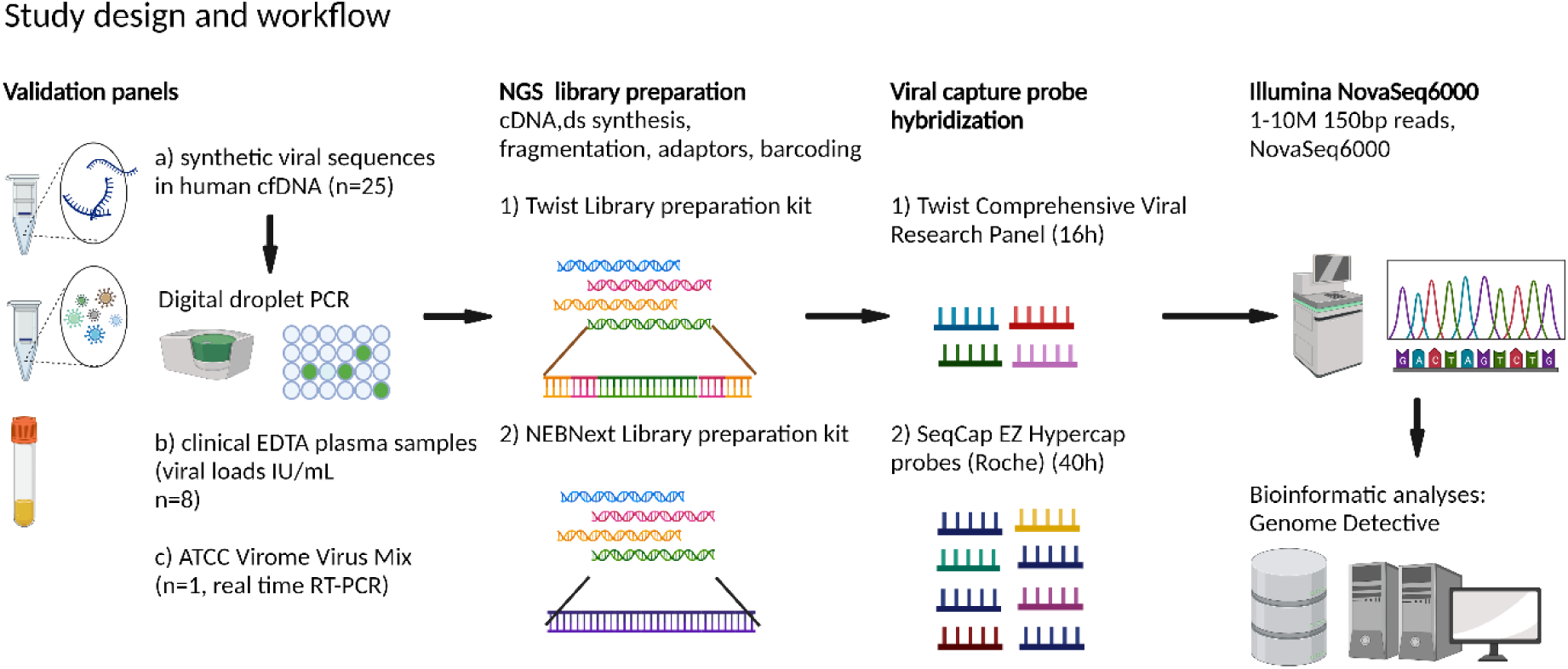
Workflows of the capture probe-based targeted metagenomic protocols compared in this study, Twist Comprehensive Viral Research, and the SeqCap EZ HyperCap (ViroCap, Roche) and, both in combination with identical bioinformatic analyses pipeline. Created using BioRender.

In order to mimic the complexity of clinical samples while reducing the number of additional viral sequences, enabling sensitivity and specificity analyses, a panel was prepared of synthetic viral sequences (10^0^ - 10^7^ copies/ml) spiked in human cell free DNA (cf DNA) background sequences (Twist Bioscience, San Francisco, USA). Synthetic viral sequences covered >99.9% of the viral genomes of SARS-CoV-2, influenza A virus (Inf A), measles, enterovirus D68, and bocavirus and were mixed with several proportions of human cfDNA (90-99.999% of weight, corresponding with proportions of up to 10-0.001%^31^ of viral nucleotides in a clinical sample^12,32,33^). Concentrations of viral sequences were determined by digital droplet PCR in triplicate (BioRad QX200). A total of 25 of synthetic mixtures were included (see **Suppl. Table 1**).

Clinical EDTA plasma samples (n=8), previously submitted to the Clinical Microbiological Laboratory for routine diagnostic testing and tested positive by qPCR^21,34^ for adenovirus (ADV), Epstein-Barr virus (EBV) and Hepatitis B virus (HBV), were included in the comparison. Viral loads ranged from 500 to 50,000 International Units (IU)/ml.

In addition, a dilution of ATCC Virome Virus Mix (MSA-2008™, ATCC, Manassas, USA) of cultivated adenovirus type F (ADV), cytomegalovirus (CMV), respiratory syncytial virus (RSV), influenza B virus, reovirus 3, and zika virus was included.

### Ethical approval

This study was approved by the medical ethics review committee Leiden/The Hague/Delft (CME number B20.002, 2020/2022).

### Nucleic acids extraction

Clinical samples and the ATCC whole virus mixture were subjected to extraction of total nucleic acids (NA) using the MagNAPure 96 DNA and Viral NA Small volume extraction kit (Roche, Basel, Switzerland) as described previously^21^.

### Viral metagenomic next-generation sequencing (mNGS)

#### Twist Comprehensive Viral Research Panel workflow

Sample preparation was performed using the Comprehensive Viral Research Panel workflow (Twist Bioscience Corp.) according to the manufacturer’s instructions. In short, 5 µl of NA was used as input for cDNA synthesis (Protoscript, New England Biolabs, Inc) followed by purification using magnetic beads, enzymatic fragmentation for 15 minutes at 30 °C, end repair and dA-tailing (Twist EF Library Prep 2.0, Twist Bioscience Corp.). Next, unique molecular identifier (UMI) adapters with unique dual barcodes (Twist UMI Adapter System, Twist Bioscience Corp.) were ligated to the fragments and amplified using PCR (12 cycles). Amplified libraries were pooled per 8 samples and library pools were used for hybridization with the Twist Comprehensive Virus probe panel, consisting of ∼1 million 120 bp probes targeting 15,488 different viral strains infecting human and animals. Hybridization was performed for 16 hours of incubation followed by several wash steps. Captured fragments were further amplified by a post-hybridization PCR (15 cycles). Finally, captured libraries were purified by a bead clean up using AmpureXP, and quantity and fragment size were determined using Qubit (Thermo Fisher, Waltham, MA, USA) and Fragment Analyser (Agilent, Santa Clara, CA, USA) respectively. Libraries were clustered and approximately 1 million 150bp paired-end reads were generated per sample, according to manufacturer’s protocols (Illumina Inc.) at GenomeScan B.V. using the NovaSeq6000.

#### SeqCap EZ HyperCap (ViroCap design, Roche)

The SeqCap EZ HyperCap workflow (Roche, Madison, USA) was performed as validated and described previously^10,21,35^. Briefly, 5 ul of NA was used as direct input (without concentration step) for enzymatic fragmentation and cDNA synthesis using the NEBNext Ultra II Directional RNA Library preparation kit V3.0 (New England Biolabs, Ipswich, MA, USA) for Illumina with several in-house adaptations to enable simultaneous detection of both DNA and RNA in a single tube per sample^12,32^. After purification, dual barcodes (NEBNext Multiplex oligos for illumina 96 unique dual index primer pairs) were attached to the fragments and amplified using PCR (21 cycles). Four barcoded samples including controls were pooled, COT (enriched for repetitive sequences) human DNA and HyperCap Universal Blocking Oligos were added before purification, following incubation for >40 hours with the SeqCap EZ HyperCap v1 (ViroCap design, 2015^14^), a collection of approximately two million oligonucleotide probes (70–120 mers) targeting all known vertebrate viruses. A complete list of the viral taxa included can be found in the supplementary tables of the manuscript by Briese et al^14^. Captured fragments were further amplified by a post-hybridization PCR (14 cycles). Finally, captured libraries were purified by bead clean up using AmpureXP, and quantity and fragment sizes were determined using Qubit (Thermo Fisher, Waltham, MA, USA) and Fragment Analyser (Agilent, Santa Clara, CA, USA), respectively. Approximately 10 million 150 bp paired-end reads were sequenced per sample according to manufacturer’s protocols (Illumina Inc.) at GenomeScan B.V. using the NovaSeq6000.

### Data analysis

#### Bioinformatic analysis

Image analysis, base calling, and quality check of sequence data were performed with the Illumina data analysis pipelines RTA3.4.4 and bcl2fastq v2.20 (Illumina). Sequence data obtained using both probe capture metagenomics methods were analyzed using a previously validated^10,21,36,37^ bioinformatics pipeline. After quality pre-processing and removal of human reads (by mapping them to the human reference genome GRCh38 (https://www.ncbi.nlm.nih.gov/assembly/GCF_000001405.26/ with Bowtie2^38^ version 2.3.4), datasets were analyzed using Genome Detective^39^ version 2.48 (accessed April – May 2023) as described previously^36^. Genome Detective includes *de novo* assembly and both nucleotide and amino acid based classification in combination with a RefSeq / Swiss-Prot Uniref database by Genome Detective^39^

Read counts were normalized for total read count and genome size using the formula: reads per kilobase per million (RPKM) = (number of reads mapped to the virus genome Y * 10^6^) / (total number of reads * length of the genome in kb)^37^. To enable analyses of the percentage of genome coverage per one million total reads, one million raw reads were randomly selected^32^ from the 10 million reads generated for the Roche protocol. The random selection from raw FASTQ files was performed with the seqtk tool (https://github.com/lh3/seqtk, version 1.3).

#### Performance metrics and statistical analyses

Sensitivity and specificity were calculated using the results from the synthetic viral sequences and the ATCC Virome Virus mix. Additional findings were considered false positives, and non-vertebrate viruses were excluded from analyses. Receiver Operating Characteristic (ROC) curves were generated by varying the number of sequence-read counts used as cut-off for defining a positive result, given a prerequisite of ≥3 genome regions covered^40^, and area under the curves (AUC) were calculated. Spearman correlations of sequence read counts with viral load, as determined by qPCR and ddPCR, were analyzed.

Limits of detection for both methods were determined using 10-fold serial dilutions of synthetic viral NA in human cfDNA background, and undiluted clinical samples. Reproducibility was determined by analyzing the coefficient of variance between runs.

Statistical analyses were performed using SPSS version 25 and 29. Statistics with P-values of 0.05 and lower were considered significant.

## Results

### Analytic sensitivity, specificity, and ROC

Detection of synthetic viral sequences in human cfDNA background, and the Virome Virus Mix, using the Twist Comprehensive Viral Research Panel and the SeqCap EZ HyperCap (ViroCap) workflow is depicted in **Suppl. Table 1**. For both methods, sensitivity was 100% (23/23, cycle threshold, C_T_, values ranging from 20 to 32). Viral target read counts ranged from 334-872,042 reads per million (RPM) for the Twist Comprehensive Viral Research workflow, and 2,171-971,610 RPM for the SeqCap EZ HyperCap workflow. Genome coverage ranged from 91.1-100% (median 99.8%), and 8.4-100% (median 97.5%), for these respective methods. Sensitivity and specificity were calculated for different thresholds for defining a positive result: i) sequence read counts and ii) genome coverage percentage, as depicted in **Figure 2** and **Table 1**. For calculation of the percentage of the viral genomes covered, a random selection of 1 million sequence reads per dataset were used. **Figure 2** shows that both RPM and genome coverage were distinctive parameters for defining a true positive result, with AUC of 99.8% for both methods when considering RPM as parameter, and ≥99.7% when considering genome coverage as parameter. Sensitivity and specificity scores of ≥95% were accomplished for both methods when 500 RPM was set as threshold, on top of a prerequisite of minimum of three distributed regions of the genome being covered (**Table 1**). Similarly, when coverage was set at 10% of the genome, both methods reached sensitivity and specificity levels of 95% and higher. Increasing the threshold for genome coverage resulted in decreased sensitivity for the SeqCap EZ HyperCap workflow, whereas it did not negatively affect the outcomes of the Twist Comprehensive Viral Research Panel workflow.

**Fig. 2.**
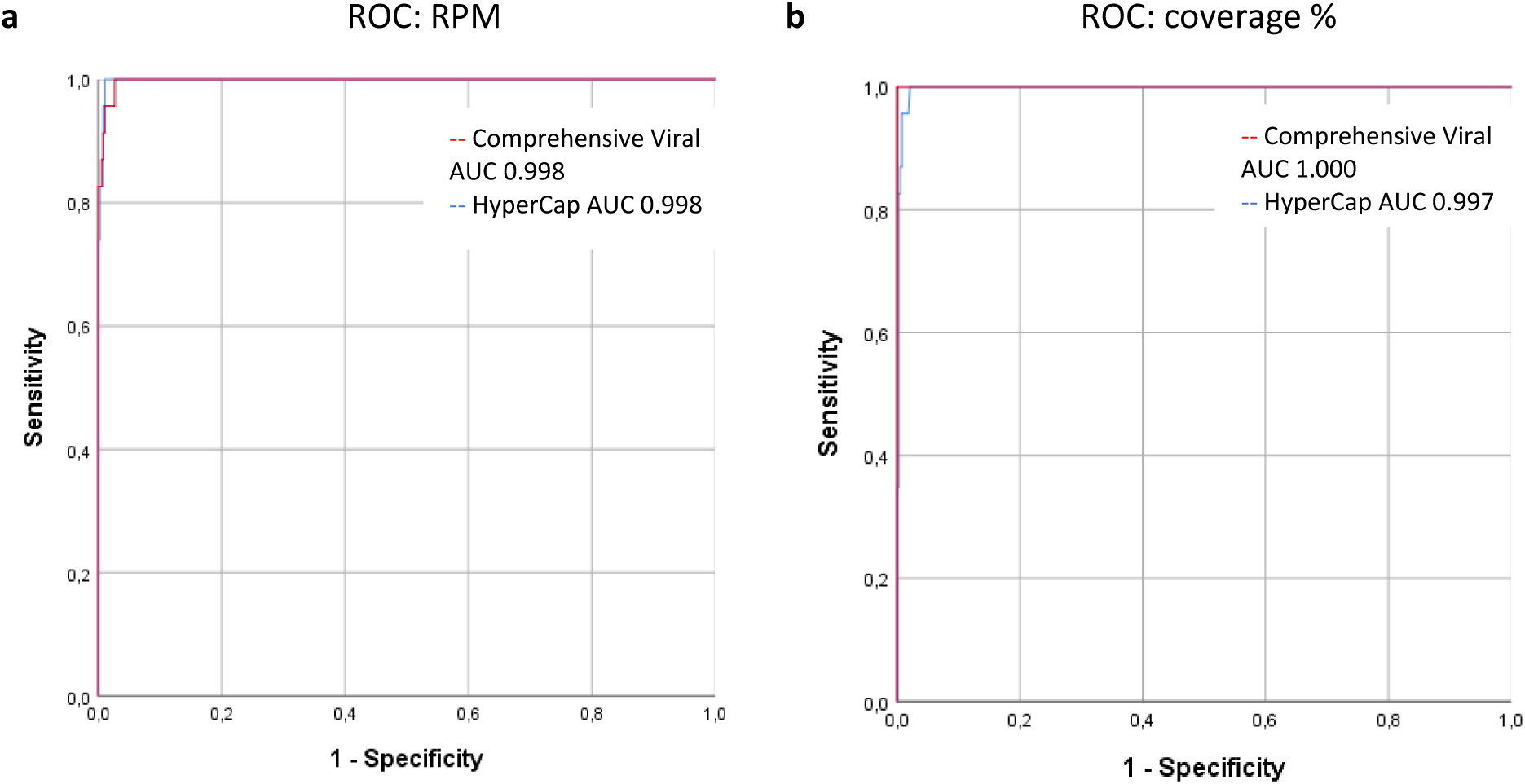
Receiver operating characteristic curves (ROC) for prediction of detection of viral sequences. using the virus capture probe based metagenomic workflows Twist Comprehensive Viral Research, and SeqCap EZ HyperCap (Roche). The validation panel consisted of synthetic viral sequences spiked in a background of human cell-free DNA (90-99.999%) and diluted ATCC Virome virus mix standard (copies/mL ranging from 10^4^ to 10^7^). **a**, ROC based on sequence read counts per million (RPM), and **b**, percentage of genome coverage, using a random selection of 1 million sequence reads per dataset. For all curves a minimum of three distributed regions of the genome covered was set as primary parameter for defining detection.

**Table 1.** Sensitivity and specificity resulting from varying thresholds based on **a**, sequence read counts and **b**, genome coverage percentage using a random selection of 1 million sequence reads per dataset, for the capture probe based metagenomic workflows SeqCap EZ HyperCap (Roche) and Twist Comprehensive Viral Research Panel workflow. Corresponding ROCs are shown in **Fig. 2**. RPM; read counts per million, LOD; limit of detection. For all tables, a minimum of three distributed regions of the genome covered was set as primary parameter for defining detection.

| Thresholds based on read counts |  |  |  |  |
| --- | --- | --- | --- | --- |
|  | 50 RPM | 500 RPM | 5,000 RPM | 50,000 RPM |
| Twist Comprehensive Viral Research |  |  |  |  |
| Sensitivity | 1.000 | 0.957 | 0.870 | 0.739 |
| Specificity | 0.960 | 0.979 | 0.995 | 1.000 |
| Corresponding LOD (c/ml)* | RNA: $10^1$<br>DNA: $10^2$ | RNA: $10^1$<br>DNA: $10^{2-3}$ | RNA: $10^1$<br>DNA: $10^{2-3}$ | RNA: $10^{2-4}$<br>DNA: $10^4$ |
| SeqCap EZ HyperCap workflow (Roche) |  |  |  |  |
| Sensitivity | 1.000 | 1.000 | 0.913 | 0.739 |
| Specificity | 0.976 | 0.984 | 0.992 | 0.997 |
| Corresponding LOD (c/ml)* | RNA viruses: $10^2$<br>DNA viruses: $10^{2-3}$ | RNA: $10^2$<br>DNA: $10^{2-3}$ | RNA: $10^2$<br>DNA: $10^{2-4}$ | RNA: $10^{2-4}$<br>DNA: $10^{4->}$ |

**b**
| Thresholds based on genome coverage |  |  |  |  |
| --- | --- | --- | --- | --- |
|  | 5% coverage | 10% coverage | 20% coverage | 90% coverage |
| Twist Comprehensive Viral Research |  |  |  |  |
| Sensitivity | 1.000 | 1.000 | 1.000 | 0.957 |
| Specificity | 0.950 | 0.968 | 0.987 | 1.000 |
| Corresponding LOD (c/ml)* | RNA viruses: $10^1$<br>DNA viruses: $10^2$ | RNA: $10^{1-2}$<br>DNA: $10^2$ | RNA: $10^2$<br>DNA: $10^2$ | RNA: $10^3$<br>DNA: $10^3-10^4$ |
| SeqCap EZ HyperCap workflow (Roche) |  |  |  |  |
| Sensitivity | 1.000 | 0.957 | 0.870 | 0.696 |
| Specificity | 0.973 | 0.981 | 0.995 | 0.997 |
| Corresponding LOD (c/ml)* | RNA: $10^2$<br>DNA: $10^{2-3}$ | RNA: $10^3-10^4$<br>DNA: $10^{2-3}$ | RNA: $10^3-10^4$<br>DNA: $10^{2-6}$ | RNA: $10^4$<br>DNA: $10^{3->4}$ |

\*Based on LOD of Inf A, SARS-CoV-2, EBV, HBV, and bocavirus. Inf A; influenza A virus, EBV, Epstein-Barr virus, HBV, hepatitis B virus.

### Correlation of viral load and sequence read counts

Viral loads as determined by ddPCR on synthetic viral sequences in human cfDNA background, and by qPCR on clinical plasma samples, were compared with sequence read counts normalized by total library size and genome size (**Figure 3**). Read counts were significantly correlated with viral loads, for both methods. Outliers were detected for samples with low viral loads, likely attributable to the stochastic effect around the limits of detection of PCR and the sequencing protocols.

**Fig. 3.**
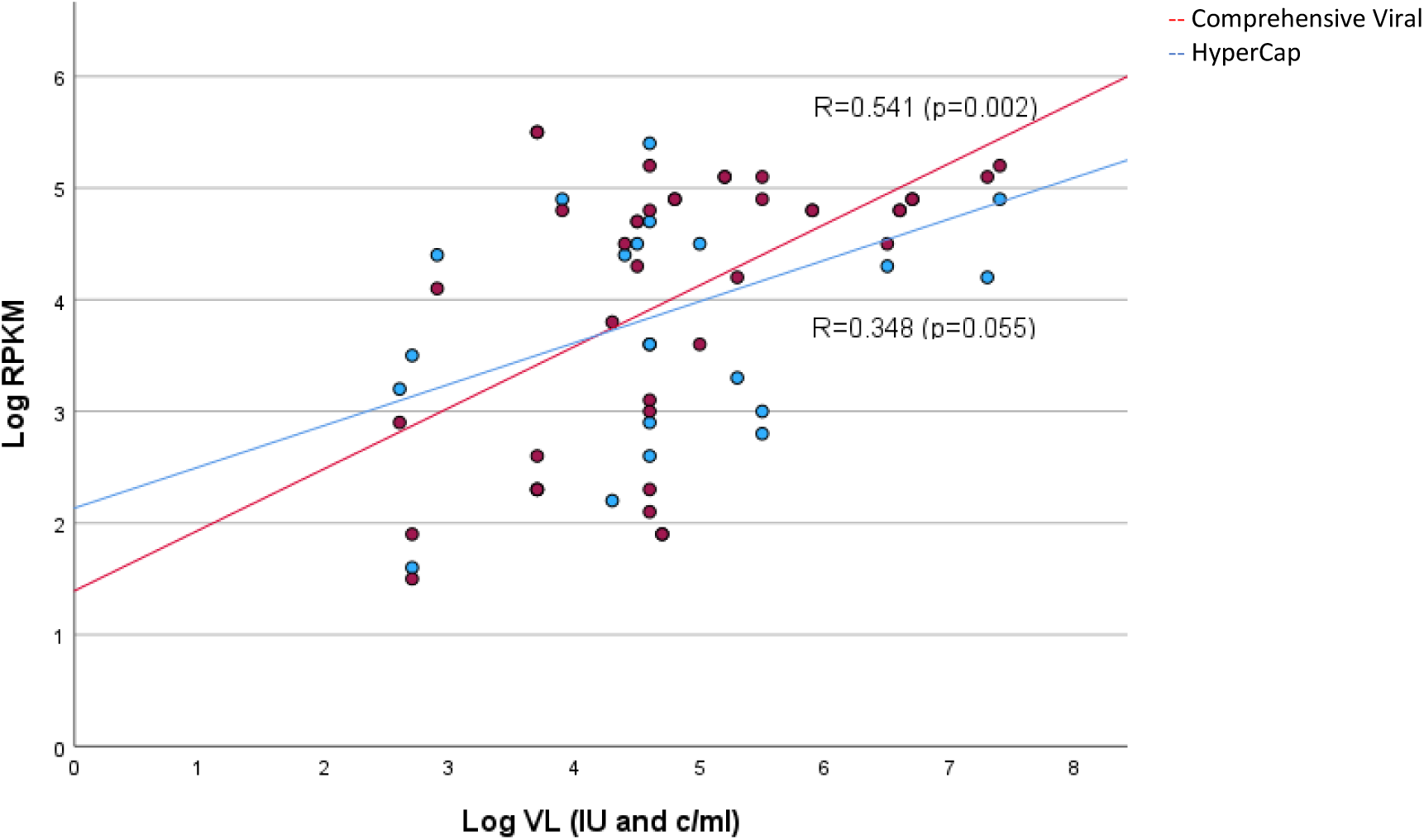
Correlation graph depicting linearity between the viral load (VL, log_10_ IU and C/ml, horizontally) and the log_10_ read counts per million per kb genome (RKPM) as generated using the virus capture probe based metagenomic workflows Twist Comprehensive Viral Research and SeqCap EZ HyperCap (Roche). Included are detections by both methods from synthetic viral sequences spiked in a background of human cell-free DNA (90/99%), dilution series (see Fig. 4), and clinical samples.

### Limits of detection

The limits of detection of both probe capture methods were analysed for several ssRNA, dsDNA, and ssDNA viruses, and is shown in **Figure 4** and **Suppl. Table 1**. The limits of detection for the RNA viruses tested were approximately 50 and 500 copies/mL for the Twist Comprehensive Viral Research Panel workflow and the SeqCap EZ HyperCap workflow, respectively. For the DNA viruses tested, the LOD was approximately 500 IU/ml for both methods, apart from the limit of detection of HBoV, which was approximately 5,000 c/ml using the SeqCap EZ HyperCap workflow.

**Fig. 4.**
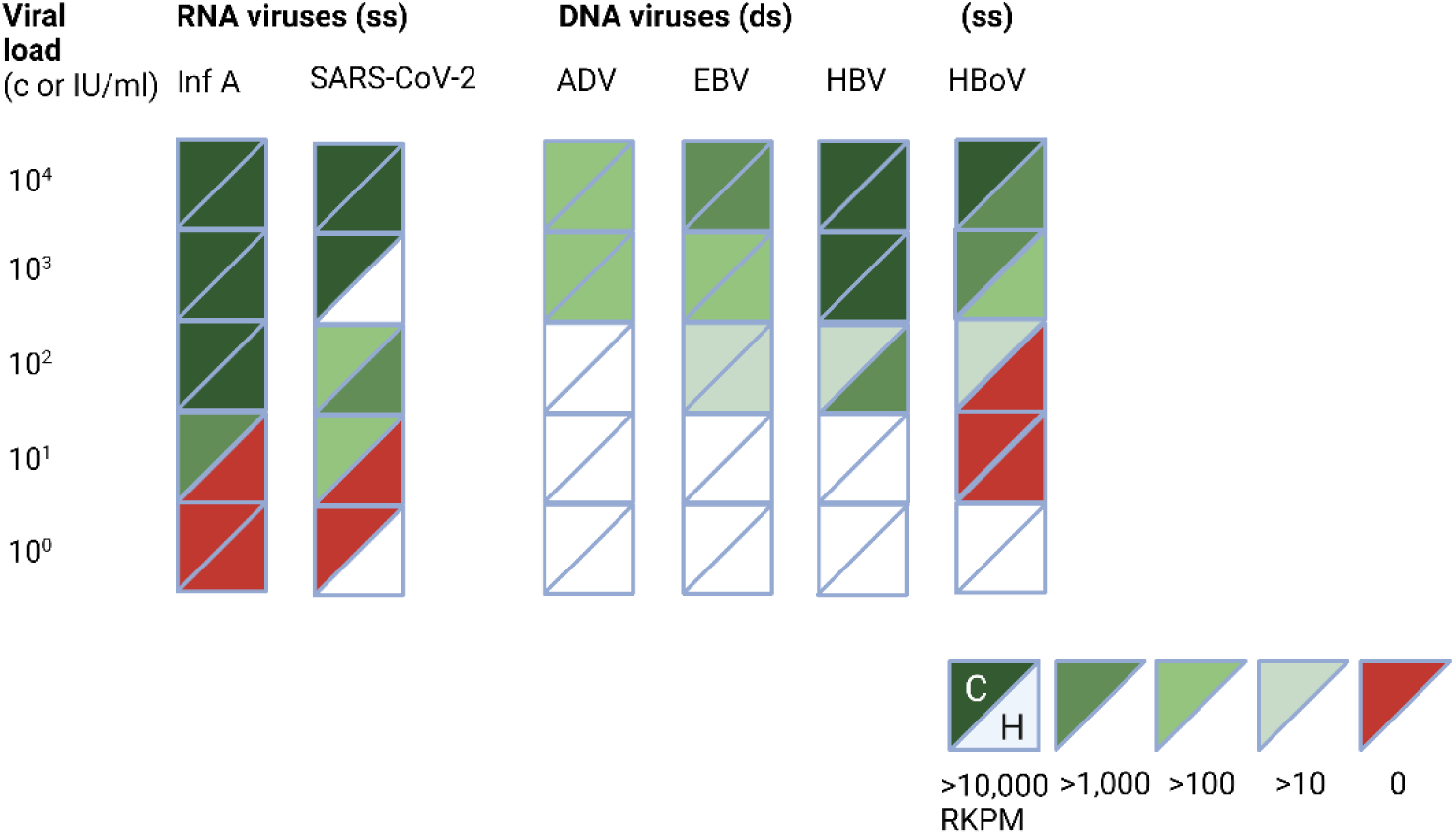
Limit of detection of viral sequences. using the virus capture probe based metagenomic workflows Twist Comprehensive Viral Research (depicted in the left upper corner, ‘C’), and SeqCap EZ HyperCap (Roche, depicted in the right lower corner, ‘H’). Read counts per million per kb genome (RPKM) are shown for different viral loads (C/ml). The samples consisted of synthetic viral sequences spiked in a background of human cell-free DNA (90-99,999%) (Inf A, SARS-CoV-2, HBoV), and clinical EDTA plasma samples (ADV, EBV, HBV). Created using BioRender.

### Reproducibility

Between-run variability as generated by both probe hybridization metagenomic workflows was studied by repeated testing of clinical samples and synthetic sequences in presence of human cfDNA background (**Figure 5** and **Suppl. Table 1**). Normalized sequence read counts and genome coverage percentage were analysed. Differences in target virus RPKM between runs were relatively low, ranging from 0.0 to 4.7% coefficients of variance.

**Fig. 5.**
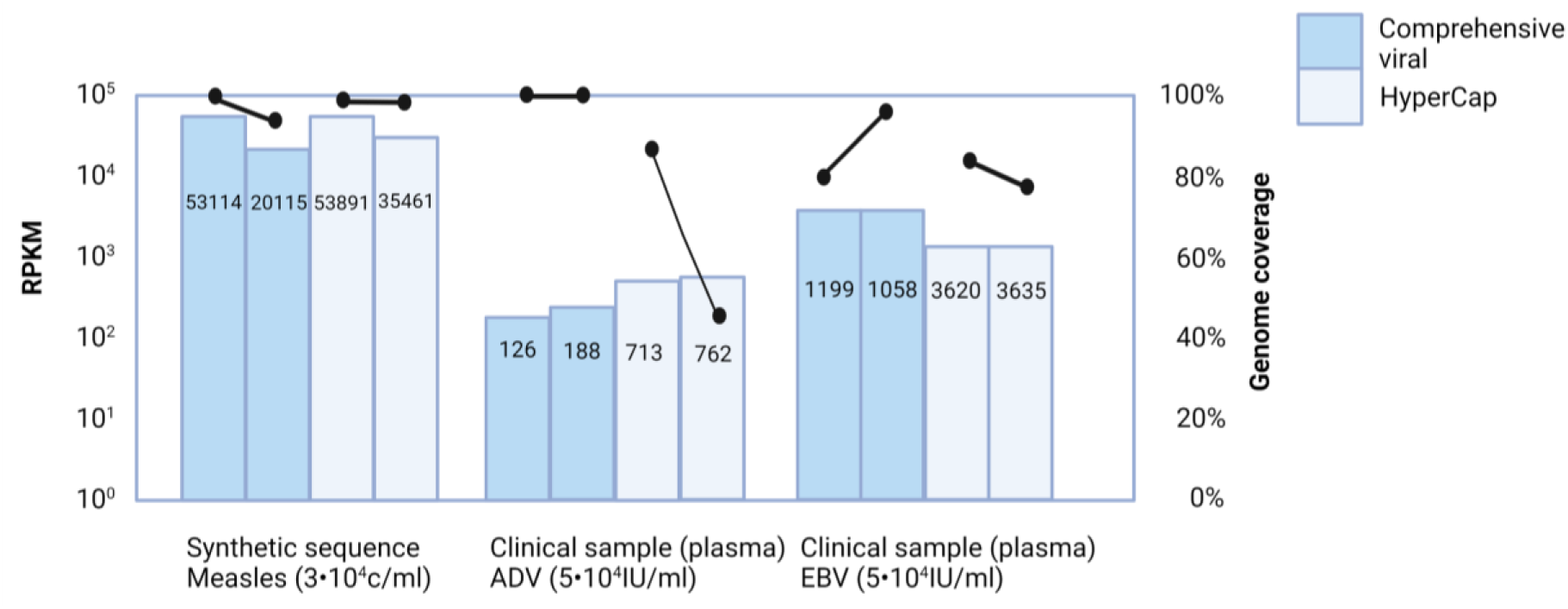
Reproducibility of read counts and genome coverage percentages. Between-run variability in RPKM (left axis) and genome coverage percentage (right axis) as generated using the virus capture probe based metagenomic workflows Twist Comprehensive Viral Research and SeqCap EZ HyperCap (Roche). Percentage of genome coverage was based on a random selection of 1 million sequence reads per dataset. Coefficients of variance in RPKM ranged from 0.0-4.7% (see **Suppl. Table 1**). Created using BioRender.

### Effect of human background sequences

The qualitative and quantitative effects of increased proportion of human background sequences on the detection of viral target sequences was studied using synthetic viral sequences spiked in a varying amount of human cfDNA background sequences (90% versus 99.999%, **Suppl. Figure 1**). No qualitative negative effect was found when the human cfDNA background proportion was increased. Quantitative target virus read counts were reduced in a single sample in which non-human reads were accounted for the largest proportion of the read count. Overall, these data indicated effective capture of target sequences.

### Application of determined thresholds to clinical samples

Optimal thresholds for defining a positive result, determined as described above (using synthetic viral sequences and the Virome Virus mixture) were applied to the eight clinical plasma samples with known viral loads (**Suppl. Table 1**). All qPCR positive findings were positive by mNGS, for both methods. Additional findings when applying a threshold of minimal 500 RPM in combination with 10% coverage of at least three regions of the genome were: torque teno viruses, adeno-associated dependoparvovirus A, and polyomaviruses. The additional findings were consistent for both methods, except for Merkel cell polyomavirus which was detected using the SeqCap EZ HyperCap workflow in two samples, indicating environmental contamination. The internal control sequences used in our laboratory, equine arteritis virus (EAV) and phocid herpes virus (PhHV, both with C_T_-values of approximately 33) were not detected and not part of the design of the Twist method, in contrast to the Roche method.

## Discussion

These data show analytical performances in ranges acceptable for clinical samples, for both probe hybridization targeted metagenomic approaches. A combination of RPM and percentage of genome coverage were optimal for defining a positive result, accompanied by sensitivity and specificity well over 95% for both methods. Limits of detection were within ranges applicable to clinical settings: 50-500 c/ml for the Twist protocol when thresholds of 500 RPM and 10% were considered. While untargeted methods are intrinsically affected by the amount of background human DNA present in (tissue) samples^41^, the results of this study show effective capturing in increasing proportions of human cell free DNA without significantly affecting the read counts and the coverage of the virus genome. This study provides the first one-to-one comparison of two pan-viral metagenomic probe capture workflows.

A recent report has studied a smaller probe panel targeting 29 human respiratory pathogenic viruses in comparison to the VirCapSeq (Roche)^42^. The authors conclude that the Twist Respiratory Virus Panel workflow was suited for detection of both respiratory co-infections and SARS-CoV-2 variants with >90% tenfold genome coverage. The latter is in line with our current data: genome coverage was generally 90-100% for samples ≥1,000 C/ml, for a range of RNA and DNA viruses. It must be noted that the required pooling of samples prior to hybridization lead to lower amounts of total reads generated for lower biomass samples. Though this potentially may result in underestimation of the performance, in practice, the sensitivity was 100% despite lower total counts in some cases using the Twist workflow. Another report was recently published on the use of the Twist Comprehensive Viral Research Panel aiming at detection of viruses involved in pediatric hepatitis cases of unknown origin, while an association with AAV2 was hypothesised^43^. In 17 cases, AAV2 was detected using targeted sequencing, while in seven of these pediatric cases AAV2 was missed by untargeted metagenomic sequencing, illustrating the significance of the use of enrichment by hybridization. With regard to cost-efficiency, a recent study compared PCR, sequence-independent single primer amplification (SISPA), and the Twist Comprehensive Viral Research Panel for the detection of Japanese encephalitis^44^. The authors concluded that the PCR panels were not able to detect all genotypes, whereas broader surveillance of vector-borne pathogens would be more effective though costly^44^. Hybridization capture has been approved by the FDA for SARS-CoV-2 variant monitoring, illustrating the acknowledged significance of this type of enrichment. The limit of detection of the SARS-CoV-2 specific hybridization method in their study was 800 copies/ml^45^, in line with our current and previous^10,46^ findings when using the broader panel. Even using the panel designed in 2015^14^ resulted in excellent genome coverage of SARS-CoV-2 due to sequence homology with animal coronaviruses and the variability in the probe design allowing for sequence mismatches^10^.

This study has several limitations. The synthetic sequences spiked in cell free human DNA did not contain other background nucleic acids such as bacterial and human RNA, though the latter proportion is generally low (<5%^47^ dependent on the sample type). Furthermore, though ssRNA, dsDNA, and ssDNA viruses were analyzed, detection and LOD results cannot be directly extrapolated to every single virus. These parameters may vary to some extent for different viruses, particularly those not included in the synthetic controls (manufactured by Twist). This was also exemplified by the lack of detection of EAV and PhHV using the Twist Comprehensive Viral Research panel. Though these viruses are not considered human pathogens, this illustrates the presence of certain restrictions with regard to the animal viruses included in the panel. Further analyses of the lists of viruses delivered by the probe designers showed that all pathogens on the WHO list of diseases with pandemic potential (https://www.who.int/news/item/21-11-2022-who-to-identify-pathogens-that-could-cause-future-outbreaks-and-pandemics) are present, in both probe panels.

To summarize, this study provides data supporting further steps towards widespread introduction of viral metagenomics for pathogen detection in clinical settings. In addition, it provides guidance for integration of probe hybridization methods in surveillance to track pathogens of pandemic potential in low biomass samples such as wastewater^48^ and wild life swabs^49^.

## Data Availability

All data produced in the present study are available upon reasonable request to the authors.

## Author’s contribution

Conceptualization: JJCV. Software: IS, Investigation; KM, AB. Methodology; KM, ECJC, JJCV. Software; IS. Formal analysis: KM, IS. Supervision; ECC, AB, SAB, JJCV. Visualization: KM, JJCV. Roles/Writing - original draft; KM, JJCV. Writing - review & editing: all authors.

