## Supplementary material for "Comparison of the performance of two targeted metagenomic virus capture probe-based methods using synthetic viral sequences and clinical samples": Suppl. Figure 1

a, Twist Comprehensive Viral Research Panel workflow

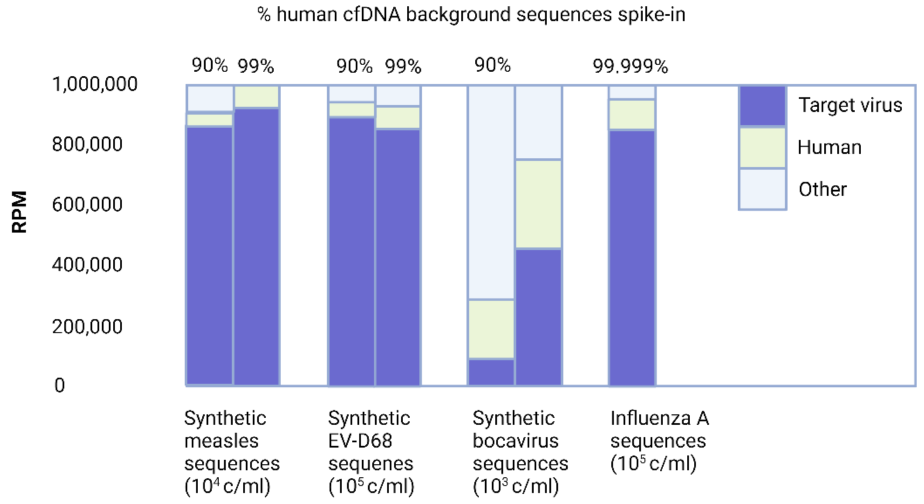


b, SeqCap EZ HyperCap workflow (Roche)

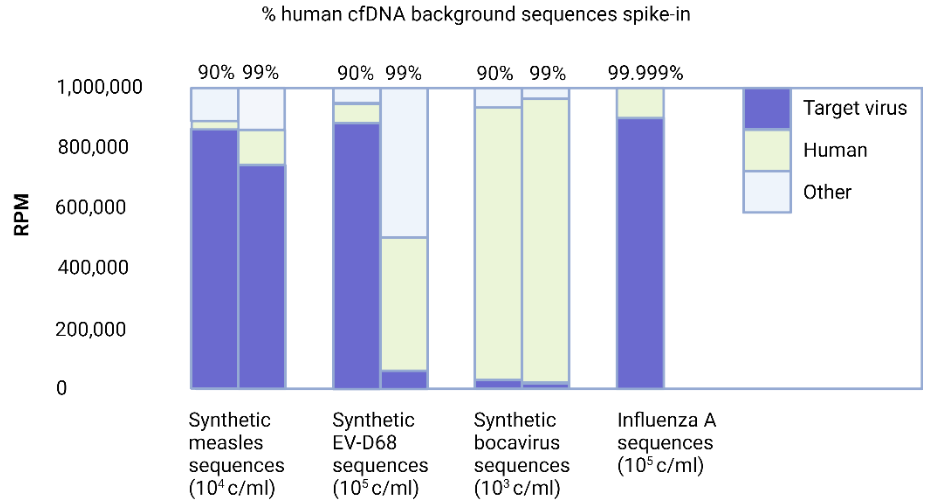


**Limited effect of spiked-in human cell free DNA background sequences (90 to 99.999%) on target virus read counts** as generated using the virus capture probe based metagenomic workflows Twist Comprehensive Viral Research workflow, and SeqCap EZ HyperCap (Roche), indicating effective capture of target sequences. Created using BioRender.
